# Performance evaluation of a multinational data platform for critical care in Asia

**DOI:** 10.1101/2021.07.10.21260243

**Authors:** Luigi Pisani, Thalha Rashan, Maryam Shamal, Aniruddha Ghose, Bharath Kumar Tirupakuzhi Vijayaraghavan, Swagata Tripathy, Diptesh Aryal, Madiha Hashmi, Basri Nor, Yen Lam Minh, Arjen M. Dondorp, Rashan Haniffa, Abi Beane

## Abstract

**Objective:** We aimed to evaluate the quality of a multinational intensive care unit (ICU) network of registries of critically ill patients established in seven Asian low and middle income countries (LMICs).

**Methods:** The Critical Care Asia federated registry platform enables ICUs to collect clinical, outcome and process data for aggregate and unit-level analysis. The evaluation used the standardised criteria of the Directory of Clinical Databases (DoCDat) and a framework for data quality assurance in medical registries. Six reviewers assessed structure, coverage, reliability and validity of the ICU registry data. Case mix and process measures on patient episodes from June to December 2020 were analysed.

**Results:** Data on 20,507 consecutive patient episodes from 97 ICUs in Afghanistan, Bangladesh, India, Malaysia, Nepal, Pakistan and Vietnam were included. The quality level achieved according to the ten prespecified DoCDat criteria was high (average score 3.4 out of 4) as was the structural and organizational performance -- comparable to ICU registries in high-income countries. Identified strengths were types of variables included, reliability of coding, data completeness and validation. Potential improvements include extension of national coverage.

**Conclusion:** The Critical Care Asia platform evaluates well using standardised frameworks for data quality and equally to registries in resource-rich settings.

**Funding:** This work was undertaken as part of the existing Wellcome Innovations Flagship award, Collaboration for Research, Improvement and Training in Critical CARE in ASIA (ref. 215522/Z/19/Z). The funder had no role in the decision to publish or in the preparation of this manuscript.

## BACKGROUND

The availability of high quality data systems to inform delivery, evaluation and improvement of health care is recognised as a central tenet of high quality health systems.^1^ In critical care, where patient populations are heterogeneous, treatments complex and where the sequelae of care requires considerable human and financial resource, intensive care unit (ICU) registries have have been instrumental in providing a mechanism for continuous, sustainable, wide scale data collection to enable service evaluation and facilitate national benchmarking of care quality. Until recently, these registries have been concentrated in high income countries, with the notable exceptions of networks in Brazil^2^ and Sri Lanka^3^. Absence of these systems in resource constrained countries severely hamper efforts to build accountability for healthcare quality.

The need to invest in systems which provide data to drive research and improvement has been highlighted by recent recommendations as part of a series of strategies to address the imbalance in quality of care that exists internationally.^1^ Recent growth in global internet connectivity and mobile technology has given opportunity for the digital health information system to be implemented and scaled in low and middle-income countries (LMICs).

The global coronavirus disease 2019 (COVID-19) pandemic has accelerated the role of registries in driving global research. For example, registries in Brazil, Australia, Europe, and in Asia have been instrumental as part of collaborations for pre-COVID-19 large scale multicentre studies,^4,5^ observational research on COVID-19^6^ and more recently interventional research, as exemplified by the randomized, embedded, multi factorial adaptive platform for community acquired pneumonia (REMAP-CAP) operational through registries in the USA and in South Asia.^6^

Whilst registries are increasingly being promoted for their role in enabling greater accountability of healthcare quality, and for their ability to facilitate multi centre clinical trials, the quality of data such systems provide requires rigorous evaluation.^7,8^ To date, evaluation of existing vertical programme assessments for digital clinical and research registries, and for the WHO endorsed district health information system platform,^9^ have focused predominantly on the ongoing challenges of missingness and inaccuracies in reporting.^10^ Few evaluations have extended to assess the timeliness, consistency, interoperability and accessibility of the data for external comparison,^11,12^ despite these dimensions of data quality being essential for clinical research.^13^

This study evaluates a network of seven federated registries operational in Asia which together use a single cloud-based platform as part of a collaboration for implementation and research in critical care. Critical Care Asia (CCA) is a collaborative programme of critical care research, training and quality improvement in Asia.^14^ The CCA currently connects 97 ICUs in seven countries to provide diverse high-quality data to generate evidence and feedback in near real time for service improvement and research, akin to the foundations of a learning health system.^15^ We sought to systematically evaluate the performance of CCA registries in Afghanistan, Bangladesh, India, Malaysia, Nepal, Pakistan and Vietnam using two pre published quality assurance frameworks. ^16,17^ We hypothesized that the quality of data arising from this federated network of registries would be high and comparable to the quality arising from ICU registries in high-resource settings.

## METHODS

### Frameworks for assessment of performance

The Directory of Clinical Databases (DoCDat) framework was established to inform researchers and clinicians on currently functioning clinical databases and to provide an independent assessment of their scope and quality.^16^ Several high quality national registries in Australia, New Zealand and in the United Kingdom have used this same framework to evaluate data quality previously.^11,12^ The framework (**Table 1**) consists of 10 items; four relating to registry coverage and six relating to reliability and validity of the data. Each item is rated on a scale of 1 to 4, with level one representing the least rigorous methods and Level 4 representing the most rigorous. The instrument was shown to have good face and content validity and to have no floor or ceiling effects.^16^ A further framework to objectively assess registry quality especially in the development and implementation phase was published in 2002 and is also used in this evaluation (**eTable 1**).^17^ This framework is divided into three main categories, and each category was applied to the central coordinating center and to the local sites.

**Table 1.** Directory of Clinical Databases (DoCDat) scoring criteria.

| Domain | Level 1 | Level 2 | Level 3 | Level 4 |
| --- | --- | --- | --- | --- |
| **A. Extent to which the eligible population is representative of the country** | No evidence or unlikely to be representative | Some evidence eligible population is representative | Good evidence eligible population is representative | Total population of country included |
| **B. Completeness of recruitment of eligible population. State when and how completeness was determined** | Few (<80%) or unknown | Some (80–89%) | Most (90–97%) | All or almost all (>97%) |
| **C. Variables included in the database** | • identifier<br>• admin info<br>• condition or intervention | • identifier<br>• admin info<br>• condition **or** intervention<br>• short term **or** long term outcome | • identifier<br>• admin info<br>• condition<br>• intervention<br>• short term **or** long term outcome<br>• major known confounders | • identifier<br>• admin info<br>• condition<br>• intervention<br>• short term outcome<br>• major known confounders<br>• long term outcome |
| **D. Completeness of data (percentage variables at least 95% complete). State when completeness was last determined:** | Few (<50%) or unknown | Some (50–79%) | Most (80–97%) | All or almost all (>97%) |
| **E. Form in which continuous data (excluding dates) are collected (percentage collected as raw data)** | Few (<70%) or unknown | Some (70–89%) | Most (90–97%) | All or almost all (>97%) or no continuous data collected |
| **F. Use of explicit definitions for variables** | None | Some (<50%) | Most (50–97%) | All or almost all (>97%) |
| **G. Use of explicit rules for deciding how variables are recorded\*** | None | Some (<50%) | Most (50–97%) | All or almost all (>97%) |
| **H. Reliability of coding of conditions and interventions. State when and how it was most recently tested:** | Not tested | Poor | Fair | Good |
| **I. Independence of observations of primary outcome** | Outcome not included or independence unknown | Observer neither independent nor blinded to intervention | Independent observer not blinded to intervention | Independent observer blinded to intervention or not necessary as objective outcome (e.g. death or lab test) |
| **J. Extent to which data are validated. State when and how it was last determined:** | No validation | Range or consistency checks | Range and consistency checks | Range and consistency checks plus external validation using alternative source |

### Performance review

Features and functions of the platform pertaining to data capture, quality and management were described and made available to a total of five reviewers. Three were independent reviewers with established track records in high quality critical care registry implementation and research in both high income settings and LMICs. Three reviewers were members of the CCA coordinating team. Independent reviewers had full access to documentation, reports, training material and platform code, pertinent to the quality assurance features of the registry.

All encounters of care reported through the seven registries during a prespecified period of six months (June-December 2020) were included. The selection of this time period enabled evaluation of established collaborating registries (Indian Registry of IntenSive care [IRIS],^18^ Pakistan registry of intensive care [PRICE]^19^ and Nepal Intensive Care Registry Foundation [NICRF]),^6^ and the inclusion of newly implemented registries (Afghanistan, Bangladesh, Malaysia and Vietnam). Basic information on these registries is detailed in **Table 2**.

**Table 2.** Characteristics of Clinical registries involved in the CCA network.

|  | All registries | Afghanistan# | Bangladesh | India | Malaysia | Nepal | Pakistan | Vietnam |
| --- | --- | --- | --- | --- | --- | --- | --- | --- |
| **Patient episodes** | **20,507** | 553 | 392 | 4,675 | 465 | 2,951 | 10,972 | 1,237 |
| **Number of ICUs** | **97** | 6 | 2 | 18 | 3 | 8 | 55 | 5 |
| **Number of beds** | **1169** | 60 | 20 | 213 | 26 | 138 | 557 | 155 |
| **Type of ICUs** |  |  |  |  |  |  |  |  |
| Mixed ICU | **33** | 5 |  | 13 | 2 | 6 | 7 | 5 |
| MICU | **19** | 1 | 1 | 1 | 1 | 1 | 12 |  |
| SICU | **20** |  |  |  |  |  | 19 |  |
| CT ICU | **1** |  |  |  |  |  | 1 |  |
| SARI ICU | **15** |  |  | 1 |  | 1 | 13 |  |
| HDU | **2** |  | 1 |  |  |  | 1 |  |
| Other | **7** |  |  | 3 |  |  | 2 |  |
| **Completeness of recruitment %** | **100** | NA | 100 | 95 | 100 | 100 | 100 | 100 |
| **Units assessing completeness, %** | **77** | 0 | 100 | 48 | 100 | 100 | 95 | 40 |
| **Long term outcomes included** | -- | no | no | yes* | no | no | yes* | no |

Data is presented as median (IQR) or n(%).
#Data collection for Afghanistan started on 2020-07-02. The remaining registries had 6 months complete collection.
Abbreviations: ICU, intensive care unit; MICU, medical ICU; SICU, surgical ICU; CT ICU, cardio-thoracic ICU; SARI, severe acute respiratory infection; HDU, high dependency unit
\*Live in some participating ICUs

### Registry structure overview

Registry structure for established registries in India, Pakistan and Nepal was already published.^15,18,20^ In brief, the CCA platform has a modular structure, where a core dataset of 33 variables captured within the first 24 hours of admission to ICU and 5 variables at discharge, provides episodic information to enable evaluation of case mix, acuity, organ support and outcomes.^18,21^ Additional modules complement the core data set providing stakeholders with a mechanism for embedding measures to evaluate care processes synonymous with care quality, and undertake observational and interventional research (**Figure 1**). The registry platform has a customisable user mobile and desktop interface and accessible data entry support tools. Minimum data connectivity requirements (3G data and offline function) along with downloadable data exports facilitate the registries adoption in settings which may previously have failed to implement digital systems due to poor internet coverage or limited access to hardware. Integrated analytics dashboards and reports displaying trends in information, activity and quality indicators provide a mechanism for service reporting and cycles of audit and feedback with the clinical teams.^15^

**Figure 1.**
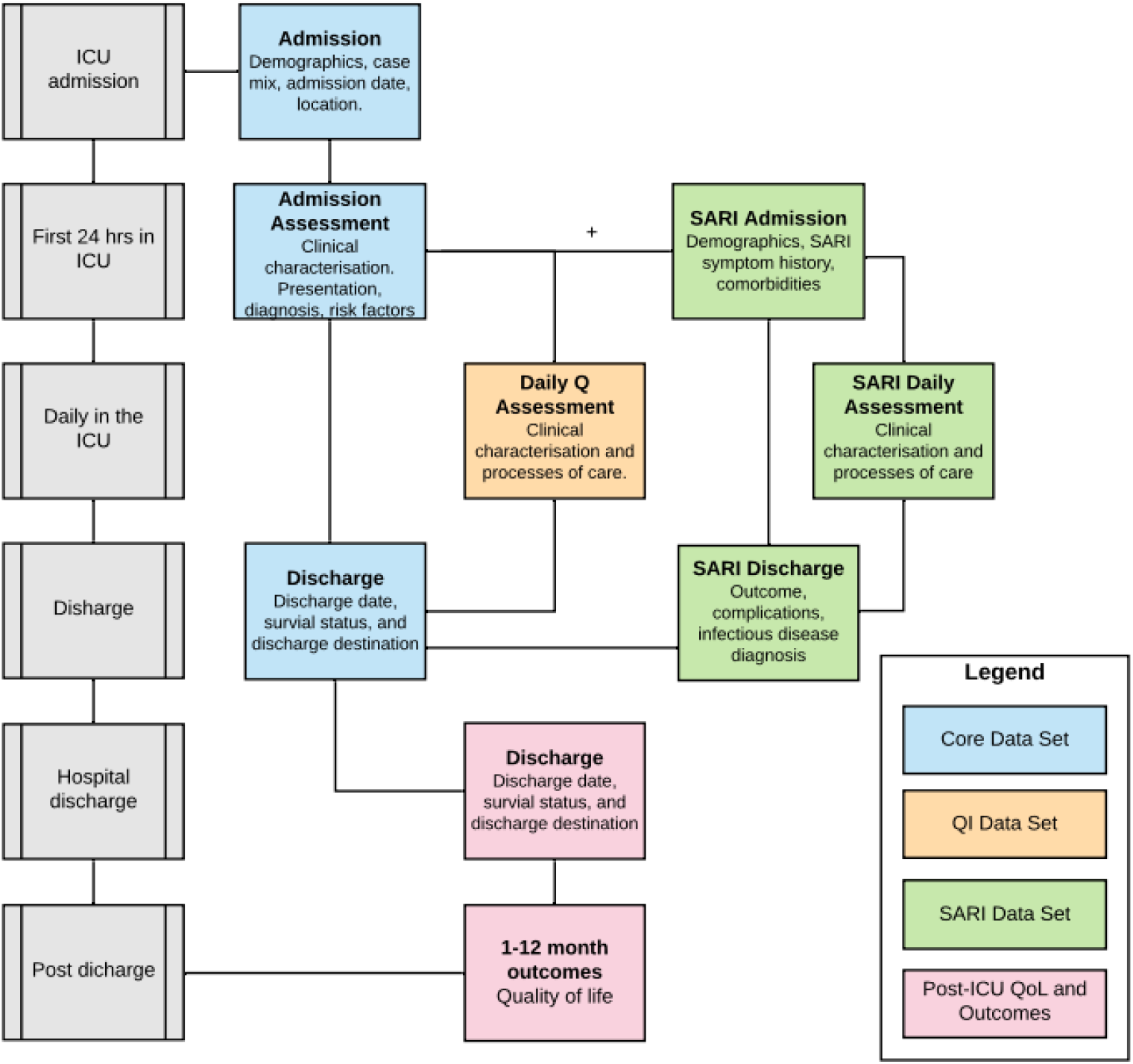
CRIT CARE ASIA registries modular data structure. Abbreviations: Q, quality; QI, quality improvement; ICU, intensive care unit (or any hospital unit involved in the project); SARI, severe acute respiratory infection; QoL, quality of life. Only the CORE data set is standard for all sites, while other data modules are optional.

The network has a federated system for registry data storage, whereby national registries house their data and are supported to establish infrastructure and skills to manage and curate data. All anonymised registry and trial data is backed up to a central server. A summary of registry implementation procedures reported using the template for intervention description and replication (TIDieR) checklist is detailed in the appendix (**Supplement file 2**).

### Data collection procedures

Data is recorded prospectively and extracted directly from patient charts by data collectors daily and contemporaneous to clinical care. Laboratory tests are reported in the ICU’s routine unit of measurement and harmonised to a single measure. A comprehensive field specification and data collection guide are made available to all stakeholders through the platform. Data collectors are remotely trained prior to commencing data collection using a demo platform and ongoing 24 hr online support is available. Follow up meetings are offered weekly to enable ongoing feedback and improvement regarding data quality and support with registry led research and audit. Census checks with independent admission data are used to monitor cohort inclusion daily or weekly at users preference. The platform’s existing internal data quality mechanisms, field completeness, value range validity and branching logic prompt users to missed or potentially spurious responses.

## RESULTS

### Assessment of performance using the DoCDat criteria

A summary of the performance of the registries using the DoCDat criteria is shown in **Table 3**, and compared to the average evaluation of other existing DoCDat databases.^11,16^ The median score achieved by the registries across all criteria was 3.4 (minimum 1.4, maximum 4). Detailed scoring of each criterion is described below, while the score assigned by each external and internal reviewer is detailed in **eTable1**.

**Table 3.** Assessment of the Crit Care Asia network registries according to the Directory of Clinical Databases (DoCDat) criteria.

|  | Crit Care Asia registries score# |  | DoCDat database* |
| --- | --- | --- | --- |
| A. Representativeness of country | 1.5 (1-2) | ■ ■ □ □ | 3 (2-4) |
| B. Completeness of recruitment | 2.7 (2-3) | ■ ■ ■ □ | 3 (1-4) |
| C. Variables included | 3.3 (3-4) | ■ ■ ■ □ | 3 (2-4) |
| D. Completeness of data | 3.8 (3-4) | ■ ■ ■ ■ | 2 (1-3) |
| E. Collection of raw data | 3.8 (3-4) | ■ ■ ■ ■ | 4 (4-4) |
| F. Explicit definitions | 4 (4-4) | ■ ■ ■ ■ | 2 (1-4) |
| G. Explicit rules | 3.8 (3-4) | ■ ■ ■ ■ | 3 (1-4) |
| H. Reliability of coding | 3.7 (2-4) | ■ ■ ■ ■ | 1 (1-4) |
| I. Independence of observations | 3.8 (3-4) | ■ ■ ■ ■ | 4 (2-4) |
| J. Data validation | 3.5 (3-4) | ■ ■ ■ □ | 3 (3-4) |

#### A. Representativeness of country

Median score 1.4. Despite the high number of ICUs in several countries, the geographic spread inside each country was limited for all registries.

#### B. Completeness of recruitment

Median score 2.6. Recruitment completeness i.e. the proportion of patients reported in the registry over the number of patients admitted to the ICU was >95% in all participating ICUs (**Table 2**). Registry team members contact each ICU on a daily or weekly basis as preferred by the registry and validate admission, discharge and bed occupancy. The recruitment completeness was assessed through a dedicated section of the online platform. The process of daily or weekly validation of recruitment completeness was conducted in all but one registry, and in 77% of all ICUs (**eTable 3**).

#### C. Variables included

Median score 3.4. All seven registries reported the core data set and were able to derive severity of illness and prediction of mortality using published scores (Acute Physiology and Chronic Health Evaluation [APACHE] II and Tropical Intensive Care Score [TropICS]).^22^ Variables included standardised diagnosis and comorbidities (Systematized nomenclature in Medicine - clinical terms [SNOMED CT] and Charlson comorbidity index), and outcomes at ICU and hospital discharge (**eTable4**). Two registries (IRIS in India and PRICE in Pakistan) also collected medium to long term patient centred outcomes (i.e. after hospital discharge) and quality of life indicators such as the Euro quality of Life 5-dimensions tool (EQ5D-3L)^23^ and scales for post traumatic stress disorders (PTSD).

#### D. Completeness of variables

Median score 3.8. All core variables were reported in the seven registries with < 5 % missingness, sustained over the 6 month period (**eTable4**). Overall, the availability of the core data set was 98.9%. All vital signs had a completeness >97%, while the variable with lowest score regarded type and dose of vasoactive drugs (96.6%).

#### E. Capture of raw variables

Median score 3.8. Raw data accounts for all fields in the core data set. Weekly meetings and 24/7 remote support between the CCA platform team and collaborating registries were reported using an online project management tool, which provided an audit trail of user queries, responses and platform development in response to recurring themes from user feedback.

#### F. and G. Explicit rules for how variables are recorded

Median scores 4.0 and 3.8 respectively. A detailed data dictionary complete with field specifications was available for all variables in the dataset and was uniform across the registries.

#### H. Reliability of coding

Median score 4.0. The CCA platform’s use of SNOMED CT and Observational Medical Outcomes Partnership (OMOP) mapping, ensures international standardized nomenclature covering both diagnostic conditions and operative procedures in all collaborating registries (**eFigure 1**). However no intra-rater or inter-rater reliability testing was performed.

#### I. Independence of observations

Median score 3.8. The primary outcome assessment for all episodes of care, was observed independent of patient care and independent from the clinical team. Similarly, secondary outcomes pertaining to vital status as 30 days-up to one year following ICU admission were captured by investigators blinded to existing encounter data.

#### J. Data validation

Median score 3.4. Data is validated internally according to the CCA dataset definitions. Fields are validated for completeness, consistency of response across sibling or parent-child fields. Inbuilt mandatory rules developed based on cycles of testing and analysis in CCA network sites ensure completeness of core dataset, and alerts within the user interface prompt users to complete supplemental fields. Illogicalities and inconsistencies in relational fields are minimised using inbuilt branching logic. Data validation reports, updated every 24hrs are accessible to end users via the platforms reports interface. Clinicians and administrators can also interrogate the CCA data set directly by downloading reports, viewing data via the real-time dashboards, or by submitting requests for analyses to the CCA registry implementation team. Free text fields are used only to supplement predetermined menus which have been generated from pre-existing guidance e.g. for Center of Disease Control definitions, or for the Acute Physiology and Chronic Evaluation (APACHE) IV diagnostic codes).

### Assessment using the framework of procedures proposed by Arts et al

The CCA platform fulfilled all criteria proposed by this framework, with the exception of 1 item (**eTable 2**) pertaining to the central coordinating center checking on interobserver variability.

## DISCUSSION

This independent evaluation of federated critical care registries from seven LMICs in Asia performed better than previously reported evaluations of multi centre databases using the DoCDat criteria.^2.11^ Key components of the platform were standardised field specification, inbuilt validation at data entry, audit reporting on completeness, consistency and validity checks of the data. The greatest limitation of the registries when evaluated against the criteria were in national geographical coverage and the absence of source verification of data.

The representativeness criteria was the lowest scoring as the CCA network spread is dishomogeneous with large differences across countries. The primary goal of capturing outcomes information is to identify high-performance hospitals or health-care delivery systems in order uncover the best practices responsible for their superior outcomes and seek to implement them in other settings. A limited coverage across the collaborating registries limits the ability to benchmark care nationally and internationally, but such benchmarking may have limited utility in healthcare systems in developing countries. This is due to both difficulty in capturing outcomes after ICU discharge and infeasibility of complex risk adjusted stratification. Although historically national coverage has been considered a key criteria to enhance data quality, we do not consider this to be the case for a federated network system spanning across several countries. The focus is on the community of practice rather than the extent of coverage, on the actual use of the data for unit level or multicenter quality improvement initiatives, audit and feedback rounds and clinical trials. Yet, efforts to increase expansion inside individual countries continue, with new centers joining the registry on a regular basis.

Some of the challenges faced by the CCA registry are specific to LMICS, others are more common and observed across registries worldwide.^11,12^ Completeness of recruitment is still not assessed in one third of the CCA ICUs and limits the exact knowledge of patients missed by the registry. On the other hand, the patient census often was higher than the reported admitted patients on ICU admission books, questioning the reliability of routinary admission books as a representation of the exact count of admitted patients. Staffing and retention of dedicated data collectors are also recognized challenges faced by registries worldwide.^11,12^ Data collection, data entry and verification are frequently carried out by staff from diverse clinical or non-clinical backgrounds with verification of data accuracy that may be seldom performed at unit level. Despite no formal audit of a sample of medical records was performed, similar rates of discrepancies (i.e. around 5%) found in previous registries^11,24^ may be expected from the CCA federated registry system. Limitations and potential flaws in reliability of registry data have been highlighted in the past.^25^ Rigorous and regular assessments of registry data such as the one performed in this article may overcome some of these limitations. Continuous audit and analysis at unit, regional and national level also contribute to strengthening data collection and interpretation procedures.

With the increased use of registries for registry-embedded clinical trials and observational research there is a drive for improved data quality.^26^ In addition to the mandatory field completeness, range checks, primitive and entity data-type constraints, additional mechanisms are in place for data quality assurance: data version management, access control for curated data sets, role-based access, verified audit trails and source verification of data. Registries can also allow a better understanding of how close standard care arms are to routine care, through the validation of trial data in the context of pre-exisiting registry data. Finally, data interoperability across multinational registries is currently being facilitated by the increasing integration of international coding systems (e.g. SNOMED), use of Common Data Models and the participation in data sharing initiatives such as the Linking of Global Intensive Care (LOGIC) consortium. ^27^

The architecture of the CCA registry facilitates ICUs retaining ownership of submitted data. The CCA registry provides contributors with a platform for capture of unit level data using a common data structure, and enables real time analysis to inform clinical care and service delivery via dashboards and collated reports. In fact, ICU beds in Asian hospitals constitute an average 9% of hospital beds, highlighting the importance of reliable and comparable data.^28^ Leveraging the same data platform, ICUs can contribute patient and hospital de-identified data to the CCA for benchmarking, multi-centre research purposes and quality improvement. Investigator initiated research can also be started by ICU registry leads within the network and on approval and agreement of clinical and institutional collaborators.

Similarly to the DoCDat criteria, Arts et al. suggested the need for transparent data definitions, standardized data collection guidelines and central training of individuals involved in data collection.^17^ The CCA failed to meet one of the suggested criteria concerning the interobserver variability checks on collected data. This would require the data collection performed by different individuals with a subsequent check against the source files, a resource-intensive procedure that constitutes a challenge for all quality clinical registries.^26^ Yet, all the other domains pertaining to both the central coordinating center and peripheral ICUs were fulfilled. This provides factual endorsement for the federated system experimented by the CCA network of multiple registries with both national and international coordination.

Across the globe, registries are now being leveraged to support large scale multi-centre clinical trials and evaluate complex improvement interventions. Regarding trial recruitment, adapted registry platforms promote rapid onboarding, inform site selection and improve patient recruitment, and can facilitate study monitoring through inbuilt data quality and validation processes.^6^ Potential limitations of registry-based trials concern the controlling for confounding and bias.^29^ The CCA network is already supporting several of the REMAP-CAP arms trials,^6,30,31^, while also enabling observational and outcome research^22^.

This study has some limitations. The assessment was limited to core data as this dataset was available throughout the all network. While other data domains will presumably share similar infrastructure scoring, the completeness of data may vary. The assessment included registries with diverse size and experience, with aggregate scoring performed without emphasis on single registries’s scores and improvement points.

## CONCLUSIONS

The CCA federated registry system is a rapidly growing network that provides high quality ICU data concerning case mix, processes of care and clinical outcomes from seven Asian countries. The system had a high performance when assessed using rigorous predefined scoring systems tackling completeness, reliability, validity and organizational infrastructure. While representativeness and interobserver reliability checks were identified as potential areas for improvement, overall performance was equal to national registries in high income settings.

## Supporting information

Supplement 2 - TiDIER checklist

Supplement 1 - additional results

## Data Availability

Data on which the scoring was performed is available upon request to CCA coordination and acceptance by individual National Registries.

## Acknowledgements

We would like to thank the registries quality independent assessors: **Jorge Salluh** (D’Or Institute for Research and Education, Rio de Janeiro, Brazil), **Dave Pilcher** (Department of Intensive Care, The Alfred Hospital, Prahran, Melbourne; Australian and New Zealand Intensive Care Research Centre, School of Public Health and Preventive Medicine, Monash University, Melbourne, VIC, Australia), **John Victor Peter** (Christian Medical College Hospital, Vellore, Tamil Nadu, India).

## Authors’ Contributions

LP, TR, AB, and RH conceived and designed the paper. MS, AG, BKTV, ST, DA, MH, BN, YLM and all collaborators collected data. TR and LP performed the statistical analysis. LP, AB and TR drafted the manuscript. RH, AMD, MS, AG, BKTV, ST, DA, MH, BN, YLM critically revised the manuscript. All authors provided final approval of the article. AB, RH, and AMD obtained funding. LP and AB take overall responsibility for the work.

## REFERENCES

1 Kruk ME, Gage AD, Arsenault C, et al. High-quality health systems in the Sustainable Development Goals era: time for a revolution. Lancet Glob Health 2018; 6: e1196–252.

2 Zampieri FG, Soares M, Borges LP, Salluh JIF, Ranzani OT. The Epimed Monitor ICU Database®: a cloud-based national registry for adult intensive care unit patients in Brazil. Rev Bras Ter Intensiva 2017; 29: 418–26.

3 Haniffa R, Mukaka M, Munasinghe SB, et al. Simplified prognostic model for critically ill patients in resource limited settings in South Asia. Crit Care Lond Engl 2017; 21: 250.

4 Soares M, Bozza FA, Angus DC, et al. Organizational characteristics, outcomes, and resource use in 78 Brazilian intensive care units: the ORCHESTRA study. Intensive Care Med 2015; 41: 2149–60.

5 Pisani L, Algera AG, Serpa Neto A, et al. Epidemiological Characteristics, Ventilator Management, and Clinical Outcome in Patients Receiving Invasive Ventilation in Intensive Care Units from 10 Asian Middle-Income Countries (PRoVENT-iMiC): An International, Multicenter, Prospective Study. Am J Trop Med Hyg 2021; published online Jan 11. DOI:10.4269/ajtmh.20-1177.

6 Aryal D, Beane A, Dondorp AM, et al. Operationalisation of the Randomized Embedded Multifactorial Adaptive Platform for COVID-19 trials in a low and lower-middle income critical care learning health system. Wellcome Open Res 2021; 6: 14.

7 Black N. High-quality clinical databases: breaking down barriers. Lancet Lond Engl 1999; 353: 1205–6.

8 World Health Organization. WHO Global diffusion of eHealth: making universal health coverage achievable. World Health Organization Geneva, 2016.

9 WHO Packages | DHIS2. https://dhis2.org/who/ (accessed May 23, 2021).

10 Hung YW, Hoxha K, Irwin BR, Law MR, Grépin KA. Using routine health information data for research in low-and middle-income countries: a systematic review. BMC Health Serv Res 2020; 20: 790.

11 Stow PJ, Hart GK, Higlett T, et al. Development and implementation of a high-quality clinical database: the Australian and New Zealand Intensive Care Society Adult Patient Database. J Crit Care 2006; 21: 133–41.

12 Harrison DA, Brady AR, Rowan K. Case mix, outcome and length of stay for admissions to adult, general critical care units in England, Wales and Northern Ireland: the Intensive Care National Audit & Research Centre Case Mix Programme Database. Crit Care Lond Engl 2004; 8: R99–111.

13 MEASURE Evaluation. Routine Health Information Systems: A Curriculum on Basic Concepts and Practice - Syllabus — MEASURE Evaluation. https://www.measureevaluation.org/resources/publications/sr-16-135a (accessed May 10, 2021).

14 CRIT CARE ASIA. Establishing a critical care network in Asia to improve care for critically ill patients in low-and middle-income countries. Crit Care Lond Engl 2020; 24: 608.

15 Beane A, De Silva AP, Athapattu PL, et al. Addressing the information deficit in global health: lessons from a digital acute care platform in Sri Lanka. BMJ Glob Health 2019; 4: e001134.

16 Black N, Payne M. Directory of clinical databases: improving and promoting their use. Qual Saf Health Care 2003; 12: 348–52.

17 Arts DGT, De Keizer NF, Scheffer G-J. Defining and improving data quality in medical registries: a literature review, case study, and generic framework. J Am Med Inform Assoc JAMIA 2002; 9: 600–11.

18 Adhikari NKJ, Arali R, Attanayake U, et al. Implementing an intensive care registry in India: preliminary results of the case-mix program and an opportunity for quality improvement and research. Wellcome Open Res 2020; 5: 182.

19 Hashmi M, Beane A, Taqi A, et al. Pakistan Registry of Intensive CarE (PRICE): Expanding a lower middle-income, clinician-designed critical care registry in South Asia. J Intensive Care Soc 2019; 20: 190–5.

20 CRIT Care Asia, Hashmi M, Beane A, Murthy S, Dondorp AM, Haniffa R. Leveraging a Cloud-Based Critical Care Registry for COVID-19 Pandemic Surveillance and Research in Low-and Middle-Income Countries. JMIR Public Health Surveill 2020; 6: e21939.

21 Hashmi M, Taqi A, Memon MI, et al. A national survey of critical care services in hospitals accredited for training in a lower-middle income country: Pakistan. J Crit Care 2020; 60: 273–8.

22 Tirupakuzhi Vijayaraghavan BK, Priyadarshini D, Rashan A, et al. Validation of a simplified risk prediction model using a cloud based critical care registry in a lower-middle income country. PloS One 2020; 15: e0244989.

23 Rabin R, Gudex C, Selai C, Herdman M. From translation to version management: a history and review of methods for the cultural adaptation of the EuroQol five-dimensional questionnaire. Value Health J Int Soc Pharmacoeconomics Outcomes Res 2014; 17: 70–6.

24 Herbert MA, Prince SL, Williams JL, Magee MJ, Mack MJ. Are unaudited records from an outcomes registry database accurate? Ann Thorac Surg 2004; 77: 1960–4; discussion 1964-1965.

25 Perner A, Bellomo R, Møller MH. Is research from databases reliable? No. Intensive Care Med 2019; 45: 115–7.

26 Litton E, Guidet B, de Lange D. National registries: Lessons learnt from quality improvement initiatives in intensive care. J Crit Care 2020; 60: 311–8.

27 Dongelmans DA, Pilcher D, Beane A, et al. Linking of global intensive care (LOGIC): An international benchmarking in critical care initiative. J Crit Care 2020; 60: 305–10.

28 Arabi YM, Phua J, Koh Y, et al. Structure, Organization, and Delivery of Critical Care in Asian ICUs. Crit Care Med 2016; 44: e940–948.

29 Frieden TR. Evidence for Health Decision Making - Beyond Randomized, Controlled Trials. N Engl J Med 2017; 377: 465–75.

30 Angus DC, Derde L, Al-Beidh F, et al. Effect of Hydrocortisone on Mortality and Organ Support in Patients With Severe COVID-19: The REMAP-CAP COVID-19 Corticosteroid Domain Randomized Clinical Trial. JAMA 2020; 324: 1317–29.

31 REMAP-CAP Investigators, Gordon AC, Mouncey PR, et al. Interleukin-6 Receptor Antagonists in Critically Ill Patients with Covid-19. N Engl J Med 2021; published online Feb 25. DOI:10.1056/NEJMoa2100433.

