## Supplement 2 - TiDIER checklist for "Performance evaluation of a multinational data platform for critical care in Asia"

***This checklist provides a structured report of the implementation process for the CCA registry.***

| TIDier Checklist | TIDier Question | CCA Platform. |
|---|---|---|
| 1. Brief | Provide the name or phrase that describes the intervention | To implement a cloud based registry platform in nine LMIC countries in Asia. |
| 2. Why | Describe any rationale, theory, or goal of the elements essential to the intervention | To provide real time data on service activity, case mix, processes of care, patient experience and outcomes for critical care services in Asia*. |
| 3. What: materials | Materials: Describe any physical or informational materials used in the intervention, including those provided to participants or used in intervention delivery or in training of intervention providers. Provide information on where the materials can be accessed (such as online appendix, URL) | Materials required for implementation included:<br>• Invitation letter<br>• CCA collaborator information<br>• Collaborator agreement (RCA)<br>• Web based portal<br>• Demo link for training User login: demoUser<br>• Password: demoUser<br>• Landscaping survey which guides platform set up and site specific requirements eg. Pakistan<br>• Institutional and ethical approvals- generic text<br>• Platform data structure<br>• Data collection guides:, QI, Core +APACHE ii SARI<br>• Medical terminology<br>• Data dictionary.<br>• Guidance for validation and data quality assurance.<br>• GCP training |
| 4. What: procedures | Procedures: Describe each of the procedures, activities, and/or processes used in the intervention, including any enabling or support activities | Please refer to supplementary table 1 below. |

| | | |
|---|---|---|
| 5. Who provided | For each category of intervention provider (such as psychologist,nursing assistant), describe their expertise, background, and any specific training given for the implementation. | ● **Project coordinators:** The project coordinator role centrally managed common scientific and administrative aspects related to the project. They assisted national coordinators in all phases of the project start-up, ethical approvals submissions, implementation and data monitoring. They assisted national and local coordinators in ensuring data safety and quality.<br>● **National coordinators***:* These individuals acted as liaison between the study team and the network sites in the country. They helped identify implementation officers and clinical leads within each country, and held an advocacy role and acted as a liaison for clinical, professional and ministerial stakeholders. They ensured that all local necessary ethical and regulatory approvals were obtained before the start of implementation.<br>● **Implementation coordinators:** Under supervision of the project coordinators, the implementation coordinators have supported the project team in each country with logistics, administration and implementation of work packages.<br>● **Site level implementation officers:** In each ICU a site level implementation officer (doctor or senior nurse) was appointed, who led local implementation, supported peer training and provided support to troubleshoot problems in the daily use of the registry. They have been reporting to the project team leads and national coordinators.<br>● **Data collectors:** Data collectors appointed in each ICU were trained alongside the clinical team in data entry and dashboard navigation as had been previously piloted in Sri Lanka and Pakistan. Implementation leads from the MORU team based in Sri Lanka closely guided this initial phase of implementation. |

| | | |
|---|---|---|
| | | ● **Data validation team:** Validation was achieved through automated rules which were tracked by the central data validation team. Checks were subsequently also performed centrally by the project team, who monitored data entry daily aided by automated scripts. Queries were sent back to the sites and addressed by the data collectors and implementation officer and overseen by the national coordinator. Daily data capture and visual display of information facilitated checks on data accuracy and completeness, these were reviewed by the implementation officer and data collector. Errors, edits and validation queries generated in the registry were stored separately, so that common errors could be identified and were acted on by the data validation team.<br>● **Development team**: A **data analyst** provided technical support for the registry and design data visualization. The **registry developer** was responsible for software development and local adaptation of the registry. A **database manager ,** along with the **database technician** covered maintenance, management and curation of the database.<br>● **Biostatistician**: They performed the statistical analyses of the observational quantitative and qualitative data, as well as analyses assessing the impact of the QI activities, including changes in ICU performance, process and outcome measures (with appropriate case mix adjustments).<br>● **Visualisation developers:** were responsible for the development of real-time data visualisation for individual ICUs and hospitals using customised dashboards supported by human-computer interaction theory. |
| 6. How? | Describe the modes of delivery (such as face to face or by some other mechanism,such as internet or telephone) of the intervention and whether it is provided individually or in a group | ● **Face to face:** Members of the NICS-MORU implementation team visited new sites to deliver training and collect feedback from stakeholders directly.<br>● **Video conferencing**: Shared screen video conferencing such as |

| | | |
|---|---|---|
| | | skype, hangouts and zoom were used to deliver training on inputting data and validation on the platform. Training for new platform features was also conducted using video conferencing.<br>● **Instant messaging**: Web-based 24 hr implementation and technical support was available via WhatsApp in all countries.<br>● **Email**: Email addresses of each country's team members and the central team were shared and email threads saved as records of discussions.<br>● **Via the registry portal**: The reporting facilities provided near real-time reporting on the data quality including completeness and census, epidemiology, severity of illness, treatment, microbiology and outcomes of ICU patients, alongside information regarding work force, unit occupancy, unit acuity, and resource utilisation. |
| 7. Where | Describe the type(s) of location(s) where the intervention occurred, including any necessary infrastructure for relevant features. | The registry implementation occured on-site in all the relevant ICUs and was centrally overseen at NICS-MORU based in Sri Lanka.<br><br>Infrastructure required for implementation has been outlined in section 4 (implementation) in detail and included:<br>● 3G connection or wifi<br>● Mobile, desktop or tablet<br>● A physical server, housed in a secure location at the network site country or at NICS-MORU's secure location. |
| 8. When and How Much? | Describe the number of times the intervention was delivered and over what period of time including the number of sessions, their schedule,and their duration, intensity, or dose | ● **Initial training and set up**: this was a period of close supervision and support including web-based 24 hr implementation and technical support available via WhatsApp. Training for the newly recruited network countries has been outlined in section 4.<br>● **Support:** Weekly video conferencing sessions with developers, researchers and users provided an opportunity for troubleshooting and feedback on feasibility, usability and enable adaptation to be discussed and prioritised.<br>● **Feedback and iteration**: Twenty four hour support via online |

| | | |
|---|---|---|
| | | messaging was available during implementation and scale up. An iterative cycle of adaptation and implementation was used to adapt the registry where necessary. |
| 9. Tailoring | If the intervention was planned to be personalised, titrated or adapted, then describe what, why, when, and how | The steps that were undertaken to ensure tailoring of the platform to each site have been outlined in detail in Section 4. They included the following:<br>● Landscaping survey to tailor the core data set and the platform identifiers- number of beds, unit names, logos, person identifiers, users and logins.<br>● The data dictionaries were produced collaboratively and local terminology as well as preferences for units of measure and bioclinical information were taken at the start.<br>● Network countries had a choice of optional forms for quality indicators and EHR features including daily observations and investigations. |
| 10. Modifications | If the intervention was modified during the course the study, describe the changes (what,why,when,and how) | ● Weekly video conferencing sessions with developers, researchers and users provided an opportunity for feedback on feasibility, usability and enabled adaptation to be discussed and prioritised.<br>● Twenty four hour support via online messaging was available during implementation and scale up. An iterative cycle of adaptation and implementation was used to adapt the registry where necessary.<br>● Examples of areas of modifications included:<br>○ **Login and access portals**<br>○ **Forms- variables/ data set -**core data set, ability to embed additional variables for research studies. Ability to embed QI audits. Ability to report pandemics/ outbreak surveillance.<br>○ **User interface:** Navigation changes<br>○ **Visualisation/ Output:** Stakeholders identified which indicators or outputs they wanted priotritsed including occupancy, acuity, equipment and resource availability, bed |

| | | |
|---|---|---|
| | | capacity<br>○ **Reports and PDF generation:** Modifications were made to what variables and indicators were reported in the unit and registry reports.<br>○ **Field labels:** These were adapted to reflect local healthcare system terminology, user interfaces were adapted to maximise data completeness and field validation and alerts were added in a personalised way to reduce user error. |
| 11. How well?: Planned | If intervention adherence or fidelity is assessed,describe how and by whom,and if any strategies were used to maintain improve fidelity, describe them | Data validation comprised automated rules incorporated in the database, on-site checks and remote checks by the central team. Automated rules flagged abnormally high or low values and missed information. The use of free text entry was restricted as much as possible.<br><br>Daily data capture and visual display of information facilitated checks on data accuracy and completeness by the implementation officer and data collector. Checks were subsequently also performed centrally by the project team, who monitored data entry daily aided by automated scripts.<br><br>Queries were sent back to the sites, which were addressed by the data collector and implementation officer and overseen by the national coordinator. Errors, edits and validation queries generated in the registry were stored separately, this allowed common errors to be identified and acted on.<br><br>Rapid feedback loops were maintained, since in the LMIC setting traceability of paper-based notes is generally limited. The use of a single platform for entering, querying, reporting and validating data increased efficiencies. |
| 12. How well?: Actual | If intervention adherence or fidelity was assessed, describe the extent to which the intervention was delivered as planned | ● **Private reporting:**<br>All ICUs had immediate access to their own units information captured through the registry via the private dashboard. This |

| | | |
|---|---|---|
|  |  | information enabled healthcare staff, administrators and researchers to evaluate trends in unit activity, severity of illness, bed occupancy, length of stay and outcomes within their respective institution. This information provided an understanding of the factors which may have been influencing care and performance.<br><br>● **Peer reporting :**<br>The aim of peer reporting was to enable evaluation of performance between comparable units. Benchmarking was used to enable stakeholders within the network to evaluate services, identify 'good or effective practice' and share this with other units. Benchmarking processes of care and outcomes (identified during the stakeholder consultation process) helped healthcare leaders identify relationships between resource, infrastructure, healthcare culture and experience, complimenting the qualitative health systems evaluation which was undertaken by the network and the training to support stakeholders to accurately evaluate and interpret the indicators of quality measured through the registry.<br><br>● **Public reporting:**<br>Anonymised aggregate data dashboards were accessible through a public portal. The aim was to provide community and public stakeholders with the opportunity to see information regarding their healthcare within their region or country promoting greater accountability of care. The dashboards were tailored to the patient and public priorities of care and included supportive information to aid interpretation. The information was used to inform those responsible for the strategic planning and financial provision of health care at regional and national level. |

Supplementary Table 1: -

| Section 4 - What? Procedures: | | | | |
|---|---|---|---|---|
| **Establishing a new registry partnership/ collaboration.** | **Site recruitment** | **Implementation** | **Validation** | **Adaptation and iteration (design-based approach)** |
| In each country a national level coordinator was appointed, to act as the liaison between the project team and the network sites in the country.<br><br>We supported the national leads to develop the team through:<br>● One-one training with the NICS Moru implementation team.<br>● Internships in established registries in the CCA network<br>● Weekly conference calls and instant messaging<br>● Monthly meetings with national coordinators<br>● Training in data collection | Stakeholders who had expressed interest in joining the registry were invited to a demo session on a shared screen using zoom, hangouts or skype.<br><br>They were then provided with access to the demo platform so that they could gain first hand experience of registry use, data sets and platform navigation.<br><br>When a new unit joined the registry a mapping exercise of the provision of ICU care was completed through a survey. The survey provided a comprehensive landscape of the ICU and directly informed registry implementation.<br><br>The survey included:<br><br>● Information regarding the facilities, equipment availability and staffing. | Frontline ICU healthcare teams, clinical stakeholders (doctors, nurses,) and AHPs received a mixture of face-to-face and remote conferencing training , in application authentication, navigation, desktop application and dashboard use.<br><br>Data collectors and site level implementation officers received training in using the platform tool and guidance on data entry. This comprised practical on-site or video conferencing training in data entry, provision of user guides, short training videos, pictorial | Data completeness, frequency of reporting and validity of data entry was reviewed on weekly calls with the NICS MORU and local implementation teams.<br><br>Data validation comprised automated rules incorporated in the database, on-site checks and remote checks by the central team. Automated rules flagged abnormally high or low values and missed information. The use of free text entry was restricted as much as possible. Daily data capture and visual display of information facilitated checks on data accuracy and completeness by the implementation officer and | An iterative cycle of adaptation and implementation was used to adapt the registry where necessary. For example, variable labels were adapted to reflect local healthcare system terminology, user interfaces were adapted to maximise data completeness and field validation or alerts were added to reduce user error.<br><br>New developments to the registry were created in the development server to ensure no disruption to the live platform's functions. These were tested by the NICS-MORU team and relevant stakeholders prior |

| | | | | |
|---|---|---|---|---|
| • Data collection and data quality was facilitated by NICS-Moru project coordinators using screen sharing and registry support documentation.<br><br>Professional bodies including critical care societies, regional training programmes, academic institutions linked to the network and, where appropriate, Ministry of Health departments, were engaged for their input and to gain support.<br><br>The approach to ethical approval for the registry was guided by the regulatory bodies within each country and informed by our experiences in undertaking collaborative projects in these sites.<br><br>Technical support for the registry, data security management, visualisation, analysis and storage was provided by the MORU team based in Sri Lanka for the duration of the funding. During the project, the team worked with local sites in each country | • The presence of unit guidelines<br>• The use of quality and safety checklists<br>• Care bundles<br>• Timings and structure of ward-rounds<br>• Handover procedures<br>• Organisational approach to care delivery, team structure and hierarchy<br><br>Team structure and roles were adapted to enable registry implementation in the different countries. Team structure reflected organisation, geography and operational structures within different ICUs.<br><br>In each ICU a site level implementation officer (doctor or senior nurse) was appointed to lead local implementation, support peer training and provide support to troubleshoot problems in the daily use of the registry.<br><br>Data collectors appointed in each ICU were trained alongside the clinical team in data entry and dashboard navigation as had been piloted in Sri Lanka.<br>An assessment of infrastructure was conducted by the NICS-MORU implementation team. Guidance was provided to help support the implementation officers, specific requirements included: | instructions and practical examples of 'use cases'.<br><br>All users were supported to complete online GCP training with oversight from the national coordinators.<br><br>Telephone support and app-based messaging groups were used to provide technical support and regular contact with the implementation officers and site coordinators and the wider clinical team for troubleshooting use case and function problems as they arose.<br><br>Problems were logged on a shared online project portal (Trello) which provided a repository of troubleshooting information, problems and solutions for evaluation following the implementation. | data collector. Checks were subsequently also performed centrally by the project team, who monitored data entry daily aided by automated scripts.<br><br>Queries were sent back to the sites, which were addressed by the data collector and implementation officer and overseen by the national coordinator. Errors, edits and validation queries generated in the registry were stored separately, so that common errors could be identified and acted on.<br><br>Rapid feedback loops were implemented, since in the LMIC setting traceability of paper-based notes is generally limited. The use of a single platform for entering, querying, reporting and validating data increased efficiency. | to deployment on the live platform.<br><br>All registry adaptation roll-outs were followed by online video conferencing training for national leads, implementation officers and other team members as required. When new features were deployed, there was a period of increased communication with implementation leads to ensure rapid problem solving.<br><br>End-users were invited to participate in semi-structured interviews and focus group discussions to evaluate the registry implementation and give feedback on the registry interface and challenges with data collection. |

| | |
|---|---|
| to build capacity in these skills to ensure local sustainability of the registry. | • 3G connection or wifi<br>• Mobile, desktop or tablet devices<br><br>The national lead was supported to assess if a physical server can be housed in the registry country. If so, the development team helped guide investment in appropriate technology which could support the registry's requirements. If a national lead chose not to store the server locally, it was housed in a secure location at NICS-MORU's secure location.<br><br>Implementation leads from the MORU team based in Sri Lanka closely guided the initial phase of implementation. Daily messaging and telephone contact from the coordinating centre supported data completeness and provided technical support. |
