## Supplement 1 - additional results for "Performance evaluation of a multinational data platform for critical care in Asia"

### SUPPLEMENTARY MATERIAL I

**eTable 1: Scoring overview of the external and internal reviewers according to the DoCDat criteria.**

| | External reviewer 1 | External reviewer 2 | External reviewer 3 | Internal reviewer 1 | Internal reviewer 2 | Internal reviewer 3 | Mean |
|---|---|---|---|---|---|---|---|
| A. Representativeness of country | 1 | 2 | 2 | 2 | 1 | 1 | 1.5 |
| B.Completeness of recruitment | 3 | 2 | 3 | 3 | 2 | 3 | 2.7 |
| C. Variables included | 3 | 3 | 3 | 3 | 4 | 4 | 3.3 |
| D. Completeness of data | 4 | 4 | 4 | 3 | 4 | 4 | 3.8 |
| E. Collection of raw data | 4 | 3 | 4 | 4 | 4 | 4 | 3.8 |
| F. Explicit definitions | 4 | 4 | 4 | 4 | 4 | 4 | 4.0 |
| G. Explicit rules | 4 | 4 | 4 | 4 | 3 | 4 | 3.8 |
| H. Reliability of coding | 4 | 4 | 2 | 4 | 4 | 4 | 3.7 |
| I. Independence of observations | 4 | 4 | 4 | 4 | 4 | 3 | 3.8 |
| J. Data validation | 4 | 4 | 4 | 3 | 3 | 3 | 3.5 |
| | | | | Overall mean | | | **3.4** |

**eTable 2. Framework of procedures for the assurance of data quality in medical registries according to Arts et al (2002).**

| CENTRAL COORDINATING CENTER | Score y/n | LOCAL SITES | Score y/n |
|---|---|---|---|
| **Prevention during set up and organization of registry** | | | |
| *At the onset of registry* | | *At the onset of participating in the registry* | |
| Compose minimum set of necessary data items | yes | Assign a contact person | yes |
| Define data and data characteristics in data dictionary | yes | Check developed software for data entry and for extraction | yes |
| Draft a data collection protocol | yes | Check reliability and completion of extraction sources | yes |
| Define pitfalls in data collection | yes | Standardize correction of data items | yes |
| Compose data checks | yes | *Continuously* | |
| Create user-friendly case record forms | yes | Train (new) data collectors | yes |
| Create quality assurance plan | yes | Motivate data collectors | yes |
| *In case of new participating sites* | yes | Make data definitions available | yes |
| Perform site visit | yes | Place data and initials on completed forms | yes |
| Train new participants | yes | Keep completed case record forms | yes |
| *Continuously* | | Data collection close to the source and as soon as possible | yes |
| Motivate participants | yes | Use the registry data for local purposes | yes |
| Communicate with local sites | yes | *In case of changes* | |
| *In case of changes (e.g. in data set)* | | Adjust forms, software, data dictionary, protocol, etc. | yes |
| Adjust forms, software, data dictionary, protocol, training material, etc. | yes | Communicate with data collectors | yes |
| Communicate with local sites | yes | | |
| **Detection during data collection** | | *Continuously* | |
| *During import of data into central database* | | Visually inspect completed forms | yes |
| Perform automatic data checks | yes | Perform automatic data checks | yes |
| *Periodically and in case of new participants* | | Check completeness of registration | yes |
| Perform site visits for data quality audit (registry data-source data) and review local data collection procedures | yes | | |
| *Periodically* | | | |
| Check interobserver and intraobserver variability | no | | |
| Perform analyses of the data | yes | | |
| **Actions for quality improvement** | | *After receiving quality reports* | |
| *After data import and data checks* | | Check detected errors | yes |
| Provide local sites with data quality reports | yes | Correct inaccurate data and fill in incomplete data | partial |
| Control local correction of data errors | yes | Resolve causes of data errors | yes |
| *After data audit or variability test* | | After receiving feedback | yes |
| Give feedback of results and recommendations | yes | Implement recommended changes | yes |
| Resolve causes of data error | yes | Communicate with personnel | yes |

**eTable 3. Completeness of recruitment by individual registry and month**

The “census” is the *weekly* comparison of the number of patients admitted to the ICU in a week against the number of patients entered in the registry.

### Afghanistan

| Month | Eligible censuses | Actually completed censuses | % | % inaccurate source data for census (reported>admitted) | Completeness of recruitment, median (reported/admitted) | IQR_25 | IQR_75 |
|---|---|---|---|---|---|---|---|
| 06/2020 | * | | | | | | |
| 07/2020 | * | | | | | | |
| 08/2020 | * | | | | | | |
| 09/2020 | * | | | | | | |
| 10/2020 | * | | | | | | |
| 11/2020 | * | | | | | | |
| 12/2020 | * | | | | | | |

***** No units were collecting census this month

**Bangladesh**

| Month | Eligible censuses | Actually completed censuses | % | % inaccurate source data for census (reported>admitted) | Completeness of recruitment, median (reported/admitted) | IQR_25 | IQR_75 |
|---|---|---|---|---|---|---|---|
| 06/2020 | 5 | 5 | 100 | 0 | 83 | 0 | 100 |
| 07/2020 | 8 | 6 | 75 | 17 | 100 | 85 | 100 |
| 08/2020 | 10 | 10 | 100 | 20 | 100 | 100 | 100 |
| 09/2020 | 8 | 8 | 100 | 25 | 100 | 100 | 101 |
| 10/2020 | 8 | 8 | 100 | 13 | 100 | 100 | 100 |
| 11/2020 | 10 | 10 | 100 | 0 | 100 | 94 | 100 |
| 12/2020 | 6 | 6 | 100 | 0 | 100 | 100 | 100 |

**India (IRIS)**

| Month | Eligible censuses | Actually completed censuses | % | % inaccurate source data for census (reported>admitted) | Completeness of recruitment, median (reported/admitted) | IQR_25 | IQR_75 |
|---|---|---|---|---|---|---|---|
| 06/2020 | 46 | 35 | 76 | 28 | 100 | 79 | 104 |
| 07/2020 | 40 | 20 | 50 | 25 | 94 | 81 | 101 |
| 08/2020 | 50 | 20 | 40 | 25 | 100 | 82 | 102 |
| 09/2020 | 43 | 23 | 53 | 21 | 100 | 87 | 100 |
| 10/2020 | 44 | 17 | 39 | 41 | 100 | 95 | 109 |
| 11/2020 | 55 | 23 | 42 | 39 | 100 | 85 | 108 |
| 12/2020 | 33 | 16 | 49 | 31 | 85 | 60 | 105 |

**Malaysia**

| Month | Eligible censuses | Actually completed censuses | % | % inaccurate source data for census (reported>admitted) | Completeness of recruitment, median (reported/admitted) | IQR_25 | IQR_75 |
|---|---|---|---|---|---|---|---|
| 06/2020 | * | | | | | | |
| 07/2020 | * | | | | | | |
| 08/2020 | * | | | | | | |
| 09/2020 | 3 | 3 | 100 | 33 | 100 | 100 | 150 |
| 10/2020 | 71 | 70 | 98 | 19 | 100 | 100 | 100 |
| 11/2020 | 90 | 87 | 97 | 6 | 100 | 100 | 100 |
| 12/2020 | 93 | 84 | 90 | 11 | 100 | 100 | 100 |

**Nepal**

| Month | Eligible censuses | Actually completed censuses | % | % inaccurate source data for census (reported>admitted) | Completeness of recruitment, median (reported/admitted) | IQR_25 | IQR_75 |
|---|---|---|---|---|---|---|---|
| 06/2020 | 92 | 92 | 100 | 26.1 | 100 | 79 | 117 |
| 07/2020 | 124 | 122 | 98 | 21 | 100 | 100 | 100 |

| | | | | | | | |
|---|---|---|---|---|---|---|---|
| 08/2020 | 124 | 124 | 100 | 20 | 100 | 100 | 100 |
| 09/2020 | 120 | 120 | 100 | 18 | 100 | 100 | 100 |
| 10/2020 | 124 | 124 | 100 | 31 | 100 | 100 | 127 |
| 11/2020 | 148 | 145 | 98 | 19 | 100 | 100 | 100 |
| 12/2020 | 248 | 248 | 100 | 16 | 100 | 100 | 100 |

**Pakistan (PRICE)**

| **Month** | **Eligible censuses** | **Actually completed censuses** | **%** | **% inaccurate source data for census (reported>admitted)** | **Completeness of recruitment, median (reported/admitted)** | **IQR_25** | **IQR_75** |
|---|---|---|---|---|---|---|---|
| 06/2020 | * | | | | | | |
| 07/2020 | * | | | | | | |
| 08/2020 | * | | | | | | |
| 09/2020 | 48 | 48 | 100 | 0 | 100 | 100 | 100 |
| 10/2020 | 241 | 235 | 97.5 | 0.4 | 100 | 100 | 100 |
| 11/2020 | 305 | 302 | 99 | 0.3 | 100 | 100 | 100 |
| 12/2020 | 183 | 183 | 100 | 0 | 100 | 100 | 100 |

**Vietnam**

| **Month** | **Eligible censuses** | **Actually completed censuses** | **%** | **% inaccurate source data for census (reported>admitted)** | **Completeness of recruitment, median (reported/admitted)** | **IQR_25** | **IQR_75** |
|---|---|---|---|---|---|---|---|
| 06/2020 | * | | | | | | |
| 07/2020 | * | | | | | | |
| 08/2020 | * | | | | | | |
| 09/2020 | * | | | | | | |
| 10/2020 | * | | | | | | |
| 11/2020 | 3 | 2 | 66 | 50 | 150 | 125 | 175 |
| 12/2020 | 6 | 0 | 0 | | | | |

### eTable 4. Completeness of data - core variables

| Form | | Variable | Availability (%) |
|---|---|---|---|
| **Admission** | 1 | Patient name | 100 |
| | 2 | Medical record number | 100 |
| | 3 | Age | 100 |
| | 4 | Gender | 100 |
| | 5 | Date of admission to hospital | 100 |
| | 6 | Time of admission to hospital | 100 |
| | 7 | Date of admission to ICU | 100 |
| | 8 | Time of admission to ICU | 100 |
| | 9 | Readmission to ICU | 100 |
| | 10 | Admission type (operative vs. non operative) | 100 |
| | 11 | Admission diagnosis | 100 |
| | 12 | Comorbidities | 100 |
| | 13 | Confirmed or suspected SARI | 99.4 |
| **Admission assessment** | 14 | Ventilatory support (mechanical vs self ventilation) | 97.6 |
| | 15 | Route of ventilatory support (ETT vs tracheostomy vs NIV) | 100 |
| | 16 | Cardiovascular support | 97.6 |
| | 17 | Type and dose of vasoactive drug | 96.6 |
| | 18 | Use of sedatives | 97.6 |
| | 19 | Use of antibiotics | 97.6 |
| | 20 | Class of antibiotic | 100 |
| | 21 | Systolic blood pressure | 97.6 |
| | 22 | Diastolic blood pressure | 97.6 |
| | 23 | Respiratory rate | 97.6 |
| | 24 | Heart rate | 97.6 |
| | 25 | Body temperature | 97.6 |
| | 26 | Renal replacement therapy | 97.5 |
| | 27 | Glasgow coma scale | 97.5 |
| **Discharge** | 28 | Date of discharge | 100 |
| | 29 | Time of discharge | 100 |
| | 30 | Discharge status | 100 |
| | 31 | Discharge destination | 100 |
| | 32 | Cardiopulmonary resuscitation during stay | 100 |
| | 33 | Withdrawal of treatment | 100 |

Abbreviations: ICU, intensive care unit; SARI, severe acute respiratory infection; ETT, endotracheal tube; NIV, non invasive ventilation.

### eFigure 1. Coding of conditions and interventions - the SNOMED CT classification scheme works

*Extracted from https://www.snomed.org/snomed-ct/five-step-briefing*

The SNOMED CT logical model defines the way in which each type of SNOMED CT component and derivative is related and represented. **The core component types in SNOMED CT are concepts, descriptions and relationships.** The model specifies how the components can be managed in an implementation setting to meet a variety of primary and secondary uses.

Every **concept** represents a unique clinical meaning, which is referenced using a unique, numeric and machine readable SNOMED CT identifier. The identifier provides an unambiguous unique reference to each concept and does not have any ascribed human interpretable meaning.

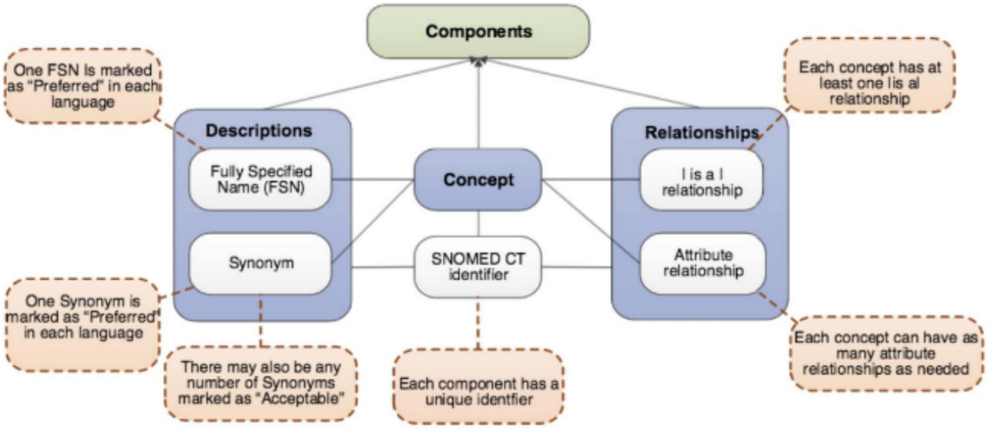

Two types of **description** are used to represent every concept – Fully Specified Name (FSN) and Synonym. The FSN represents a unique, unambiguous description of a concept's meaning. This is particularly useful when different concepts are referred to by the same commonly used word or phrase. Each concept can have only one FSN in each language or dialect. A synonym represents a term that can be used to display or select a concept. A concept may have several synonyms. This allows users of SNOMED CT to use the terms they prefer to refer to a specific clinical meaning.

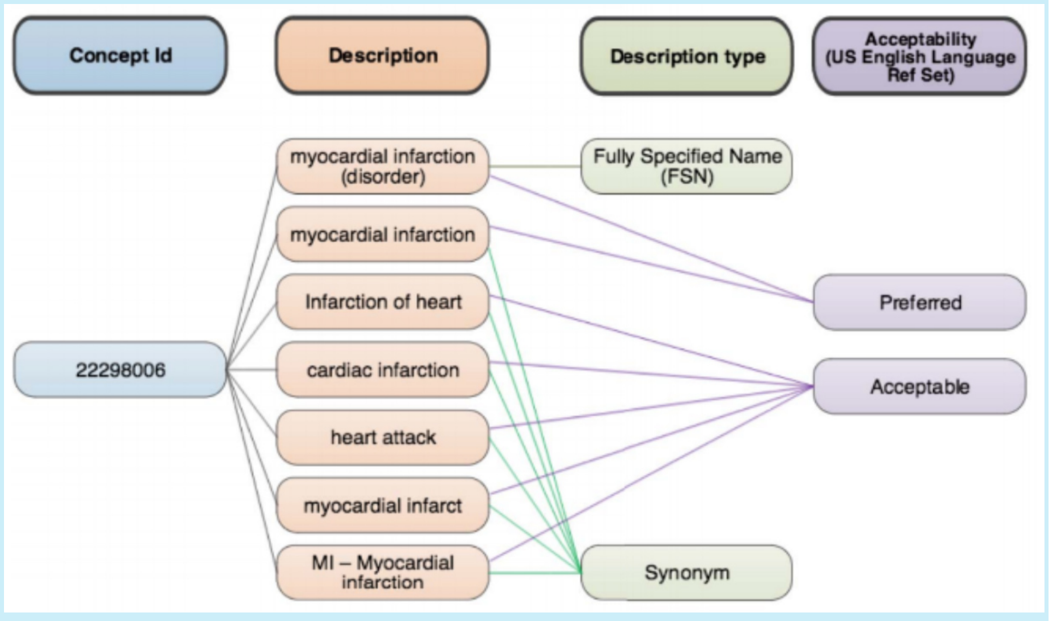

A **relationship** represents an association between two concepts. Relationships are used to logically define the meaning of a concept in a way that can be processed by a computer. A third concept, called a relationship type (or attribute), is used to represent the meaning of the association between the source and destination concepts. There are different types of relationships available within SNOMED CT.

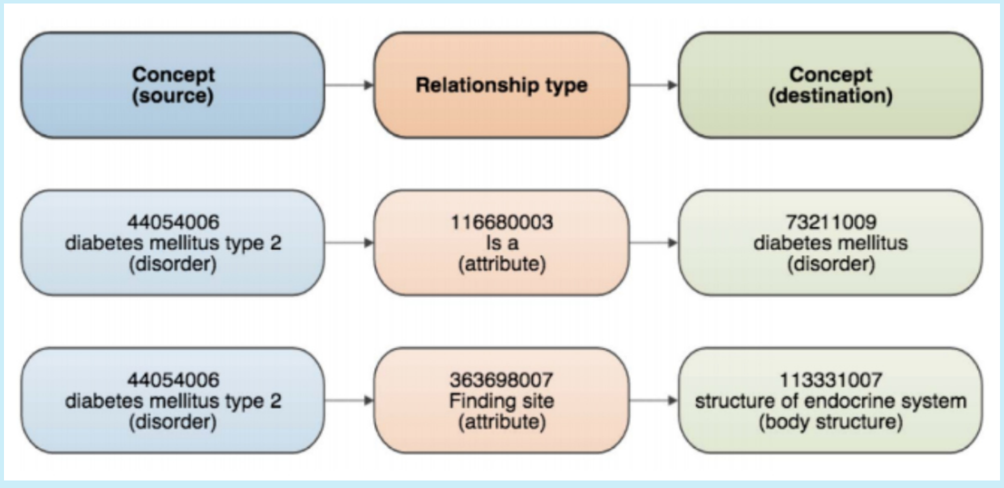
